# Comorbid hypertension and its associated factors among diabetic outpatients in West Shewa Zone public hospitals, Ethiopia: an institution based cross-sectional study

**DOI:** 10.1101/2023.03.25.23287734

**Authors:** Delessa Hirpa, Daba Abdissa

**Author notes:** **Corresponding author** Delessa Hirpa. Email address Delessa Hirpa. Email address Daba Abdissa.

## Abstract

**Objectives:** Hypertension (HTN) is the main contributor to the worldwide burden of disease and it is frequently coexists with diabetes and exacerbates its complications. The purpose of this study was to evaluate the prevalence and determinants of HTN among diabetic outpatients at West Shoa Zone public hospitals.

**Methods:** Facility based cross-sectional study was conducted from June to December, 2020 among diabetic patients attending their follow up at West Shewa public hospitals, Ethiopia. Data were collected using interviewer administered pretested structured questionnaire. A variable having a p-value of <0.25 in the bivariable analysis were subjected to multivariable analysis to avoid confounding variable’s effect. Adjusted odds ratios were calculated at 95% confidence interval and considered significant with a p-value of ≤ 0.05.

**Results:** A total of 390 participants were included in the study. Their mean age was 46.45 years (±15.6) years. Our study found that there was high prevalence of hypertension among diabetic patients. Age ≥50 year, obesity, family history of hypertension and being single were associated with hypertension among participants. Hence, necessary actions are recommended by responsible bodies for identified problems.

## Introduction

Hypertension (HTN) is the major contributor to the worldwide burden of disease and to global mortality (1) and it is forecast to reach 1.56 billion worldwide by 2025 (2). Diabetes mellitus (DM) and HTN are interrelated disorders, each powerfully predisposing to the development of each other and HTN is common problem in diabetics. Studies have shown that the expected rise in HTN is also related to the scourge in the prevalence of DM (3,4).

HTN is estimated to cause 7.5 million deaths each year, accounting for 57 million disability-adjusted life years and accounts for about 6% of deaths worldwide (5,6). Globally, approximately one billion people have HTN of these, two-thirds are in developing countries (7). Globally, about one in every four men and one in every five women have HTN (8). HTN leads to a number of disabling complications (4).

Despite being preventable disease, DM and HTN fall among top 10 leading causes of death globally(9). HTN, among DM patients, is a global public-health challenge and its frequency among the DM patients is almost twice of the non-diabetic patients (10,11). Compared with other cardiovascular disorders, HTN is the most common comorbid disease in diabetic patients and its consequences are devastating if not controlled (12,13).

The coexistence of HTN and DM is a major contributor to the development and progression of DM complications (14). Up to 80% of people with DM will die of cardiovascular disease (14,15). The comorbid HTN in DM patients is attributed to the risk of death and cardiovascular disease (CVD) events by 44% and 41%, respectively, as compared to 7% and 9% of the these risks in people with DM alone (16). Furthermore, the development of HTN in DM patients increases healthcare costs and complicates treatment strategy (17). DM approximately doubles risk of CVD and concomitant HTN nearly doubles that risk again. Moreover, DM related renal dysfunction, and increased arterial stiffness have been proposed as contributing factors for the development of HTN in diabetics (18).

Focusing on detecting and managing HTN in patients with DM is one of the most effective things that can be done to prevent DM complications. Overweight/obesity, older age, lack of physical exercise, unhealthy diet, smoking and family history of HTN are major risk factors for HTN development among DM patients (19,20).

As per authors knowledge there are not enough studies in Ethiopia regarding the prevalence and associated factors of HTN among DM patients. Furthermore, there is no evidence available in the study area on this issue. Hence, this study was tried to solve this gap which aims to reduce the incidence and mortality associated with this disease.

## Main text

### Study setting, design and period

An institution based cross-sectional study conducted from June to December, 2020 among 390 diabetic patients attending their follow-up at chronic illness clinic of West Shewa public hospitals, Ethiopia. According to the 2007 census of Ethiopia, the total population of the West Shoa Zone is estimated to be 2,058,676 of which 1,028,501 are males and 1,030,175 are females in 2018/2019. In this zone, there were 520 health posts, 92 health centers, and 8 hospitals. These Hospitals were ambo referral Hospital, Ambo general hospitals, Gendeberet general hospital, Bako Primary hospital, Jaldu Primary hospital, Enchini Primary hospital Gudar Primary hospital and Gedo general hospitals.

### Eligibility criteria

All patients with DM age ≥ 18 years on follow-up at the study area and gave consent to participate were included in the study and patients with a severe cognitive, or hearing impairment, who were critically ill and pregnant women were excluded from the study.

### Sample size determination

The sample size was calculated using a single population proportion formula with 95% confidence interval, 37.4% proportion and a margin of error 5%(21) i.e. 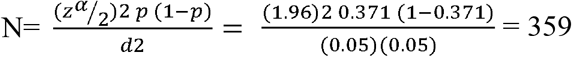 and by adding 10% for nonresponse the final sample size was 396.

### Sampling technique and procedure

Ambo University Referral Hospital was selected purposively, and the other three hospitals (Ambo General Hospital, Gedo General Hospital and Guder general hospital) were selected randomly. Then, the sample was proportionally allocated for each hospital. The study subject from each selected hospitals was taken by systematic random sampling. Six participants were excluded because of incomplete Laboratory investigations (Table 1).

**Table 1:** Proportional allocation of sample size to randomly selected hospitals, 2020

| Selected hospitals | Number of DM population | Proportionally allocated Samples |
| --- | --- | --- |
| Ambo University referral hospital | 1500 | 138 |
| Ambo general hospital | 1200 | 110 |
| Guder primary hospital | 600 | 56 |
| Gedo general hospital | 1000 | 92 |
| Total | 4300 | 396 |

### Data collection tool and procedure

Data were collected by using a structured interviewer administered questionnaire which was developed by researchers from relevant literatures. Behavioral variables were assessed based on WHO Step wise approach for chronic disease risk factor surveillance (22). Clinical variables were taken from patient record review and physical measurements were conducted.

Body weight was measured to an accuracy of 0.1□kg by using portable weight scale machine. Subjects were barefoot and wearing light indoor clothing. Height was measured using a calibrated stadiometer to the nearest 0.1 cm. All measurements were repeated two times for accuracy. Body mass index (BMI) was calculated as the ratio of weight in kilograms (Kg) to the square of height in meters (m^2^).

Blood pressure (BP) were measured from left arm at level of the heart using mercury-based sphygmomanometer after the participants rested for few minutes before their blood pressure was measured. All three measurements were performed in a sitting position for at least five minutes apart.

#### Biochemical Measurements

Blood sampling consisted of drawing five ml of blood under aseptic conditions using plain vacationer tubes will be obtained after an overnight fast (≥ 8hrs). The blood samples will left at room temperature to allow clotting for 15-20 minutes and centrifuged at 3000 rpm for 10 minutes. The levels of total cholesterol (TC), glucose and triglycerides (TG) measured by COBAS c 311 chemistry analyzer (Roche diagnostic Germany). All laboratory measurements were done as per guideline. Four BSC nurses were collected the data with supervision of principal investigator and 2 supervisors.

### Operational definition

#### Hypertension

In our study context a study participant was classified as hypertensive if the average SBP/DBP was ≥140/90mmHg and/or or patients on antihypertensive therapy.

#### Critically ill

Patients who are unable to communicate and abnormal conscious

### Data entry, processing and Statistical Analysis

Data were categorized, cleared, compiled and coded, checked for completeness, accuracy then entered into Epi data version 3.1 and then exported to SPSS. Both bivariable and multivariable logistic regression analysis was done and variables that were significant in bivariable with a p-value of <0.25 were retained for further consideration with multivariable logistic regression to control confounders. Finally significance of statistical association was assured using 95% confidence interval and a p-value of (<0.05) was considered significant in multivariable regression. The necessary assumption of model fitness during logistic regression was checked using Hosmer-Lemeshow goodness-of-fit test statistics. Multicollinearity was checked by a variable inflation factor and showed no multicollinearity.

### Data quality control

Before the actual data collection two day intensive training about the sampling procedures and aim of the study was given to data collectors by the principal investigator. The tool was pre-tested on 5% of the sample size in nearby Holeta Hospital and all required revisions were made to the study tool based on the pre-test. Experienced enumerators were recruited for the data collection and selected participants were oriented about the study. Continuous follow up and supervision was made by the 2 supervisors and principal investigator and collected data was reviewed and checked daily for clarity and completeness.

Standard operating procedure was used for all laboratory analysis of blood samples. The tests were conducted based on the manufacturers’ instruction. The quality assurance principles for pre-analytical, analytical and post-analytical stages were applied to assure the quality result. Those intermediate results were repeatedly checked. Visual inspections of neatness of the lab and working bench performed to avoid cross contamination. There was properly recording of the daily result and daily follow up by principal investigator.

## Results

### Socio-demographic characteristics of respondents

During the study period, 390 diabetic patients participated in the study. The mean age of the participants was 46.45 years (±15.6). Almost half of the respondents were males (50.8%), more than three-fourth was married (76.4%) and majority of participants was urban dwellers (62.3%) (Table2).

**Table 2:** Socio-demographic characteristics of patients and prevalence of HTN of participants with diabetes mellitus, 2020

| Variables | Category | Number | Percent | HTN status |  |
| --- | --- | --- | --- | --- | --- |
|  |  |  |  | Yes (%) | No (%) |
| Sex | Male | 198 | 50.8 | 58 (70.7%) | 140 (29.3%) |
|  | Female | 192 | 49.2 | 109 (56.8%) | 83 (43.2%) |
| Age (Year) | Mean (SD) | 46.45(15.67) |  |  |  |
|  | Under 30 | 70 | 17.7 | 24 (34.3%) | 46 (65.7%) |
|  | 30-39 | 59 | 15.1 | 23 (39.0%) | 36 (61.0%) |
|  | 40-49 | 97 | 24.9 | 49 (50.5%) | 48 (49.5%) |
|  | 50 and above | 164 | 42.1 | 96 (58.5%) | 68 (41.5%) |
| Average monthly income (ETB) | <1000 | 87 | 22.3 | 54 (62.1%) | 33 (37.9%) |
|  | 2000-2999 | 27 | 6.9 | 19 (70.4%) | 8 (29.6%) |
|  | >3000 | 193 | 49.5 | 126 (65.3%) | 67 (34.7%) |
| Marital status | Married | 298 | 76.4 | 197 (66.1%) | 101 (33.9%) |
|  | Single | 73 | 18.7 | 41 (56.2%) | 32 (43.8%) |
|  | Divorced | 13 | 3.3 | 6 (46.2%) | 7 (53.8%) |
|  | Widowed | 6 | 1.5 | 5 (83.3%) | 1 (16.7%) |
| Religion | Orthodox | 199 | 51.0 | 128 (64.3%) | 71 (35.7%) |
|  | Protestant | 154 | 39.5 | 100 (64.9%) | 54 (35.1%) |
|  | Muslim | 22 | 5.6 | 14(63.6%) | 8 (36.4%) |
|  | Wakefata | 12 | 3.1 | 5(41.7%) | 7 (58.3%) |
|  | Others | 3 | .8 | 2 (66.7%) | 1(33.3%) |
| Educational status | Can't read and write | 85 | 21.8 | 58 (68.2%) | 27 (31.8%) |
|  | Only Read & Write | 28 | 7.2 | 22 (78.6%) | 6(21.4%) |
|  | primary education (1-8) | 93 | 23.8 | 56 (60.2%) | 37 (39.8%) |
|  | Secondary education (9- | 70 | 17.9 | 43(61.4%) | 27 (38.6%) |
|  | 12) |  |  |  |  |
|  | Level I-IV and Diploma | 52 | 13.3 | 32 (61.5%) | 20 (38.5%) |
|  | Degree and above | 62 | 15.9 | 38 (61.3%) | 24 (38.7%) |
| Occupational status | House wife | 55 | 14.1 | 32 (58.2%) | 23 (41.8%) |
|  | Gov't Employee | 123 | 31.5 | 72 (58.5%) | 51 (41.5%) |
|  | Non-Gov't Employee | 32 | 8.2 | 22 (68.8%) | 10 (31.3%) |
|  | Student | 33 | 8.5 | 16 (48.5%) | 17 (51.5%) |
|  | Merchant | 41 | 10.5 | 26 (63.4%) | 15 (36.6%) |
|  | Daily laborer | 11 | 2.8 | 8 (72.7%) | 3 (27.3%) |
|  | Farmer | 95 | 24.4 | 73 (76.8%) | 22 (23.2%) |
| Residence | Rural | 147 | 37.7 | 98 (66.7%) | 49 (33.3%) |
|  | Urban | 243 | 62.3 | 151 (62.1%) | 92 (37.9%) |
| Family History of Hypertension | Yes | 83 | 21.3 | 58 (69.9%) | 25 (30.1%) |
|  | No | 307 | 78.7 | 191 (62.2%) | 116 (37.8%) |
| Family History of DM | Yes | 105 | 26.9 | 64 (61.0%) | 41 (39.0%) |
|  | No | 285 | 73.1 | 185 (64.9%) | 100 (35.1%) |

### Clinical and behavioral characteristics of participants

Among study participants, majority of them was diagnosed with diabetes for less than 10 years. A total of 201(51.5%) study participants were in the normal category of BMI and more than half (56.9%) used a noninsulin drug (Table 3).

**Table 3:** Clinical and behavioral characteristics of patients and prevalence of HTN of participants with diabetes mellitus, 2020.

| Variables | Category | Number | Percent | Outcome of HTN |  |
| --- | --- | --- | --- | --- | --- |
|  |  |  |  | Yes (%) | No (%) |
| Diabetes Mellitus Type | T1DM | 111 | 28.5 | 66 (59.5%) | 45 (40.5%) |
|  | T2DM | 279 | 71.5 | 183 (65.6%) | 96 (34.4%) |
| Duration of DM | <5yrs | 207 | 53.1 | 130 (62.8%) | 77 (37.2%) |
|  | 5-10yrs | 107 | 27.4 | 68(62.6%) | 40 (37.4%) |
|  | ≥10yrs | 76 | 19.5 | 52 (68.4%) | 24 (31.6%) |
| Treatment regimen | Oral hypoglycemic agents | 222 | 56.9 | 142 (64.0%) | 80(36.0%) |
|  | Injection (Insulin) | 129 | 33.1 | 83 (64.3%) | 46(35.7%) |
|  | Oral and injection | 39 | 10.0 | 24 (61.5%) | 15 (38.5%) |
| Statin Treatment | Yes | 37 | 9.5 | 22 (59.5%) | 15 (40.5%) |
|  | No | 353 | 90.5 | 227 (64.3%) | 126 (35.7%) |
| BMI (kg/m2) | Low (<18.5) | 22 | 5.6 | 16 (35.7%) | 6 (27.3%) |
|  | Normal (18.5-24.9) | 201 | 51.5 | 124(61.7%) | 77 (38.3%) |
|  | Overweight (25-29.9) | 126 | 32.3 | 82 (65.1%) | 44 (34.9%) |
|  | Obese (≥30) | 41 | 10.5 | 27 (65.9%) | 14 (34.1%) |
| Hypertension | Yes | 169 | 43.3 | 117 (69.2%) | 52 (30.8%) |
|  | No | 221 | 56.7 | 132 (59.7%) | 89 (40.3%) |
| Alcohol intake | Yes | 133 | 34.1 | 80 (60.2%) | 53 (39.8%) |
|  | No | 257 | 65.9 | 169 (65.8%) | 88 (34.2%) |
| Smoking status | Yes | 47 | 12.1 | 32 (68.1%) | 15 (31.9%) |
|  | No | 343 | 87.9 | 162 (47.2%) | 181 (52.8%) |
| Vigorous-intensity aerobic physical activity | Yes (≥75-150 minutes/week) | 118 | 30.3 | 77 (65.3%) | 41 (34.7%) |
|  | No (<75-150 minutes/week) | 272 | 69.7 | 172 (65.1%) | 100 (36.8%) |
| Moderate-intensity aerobic physical activity | Yes(≥150- 300 minutes/week) | 141 | 36.2 | 97 (61.7%) | 54 (38.3%) |
|  | No (<150-300 minutes/week) | 249 | 63.8 | 162 (65.1%) | 87 (34.9%) |
| FBS (mg/dl) | Normal (<126 mg/dl) | 269 | 69.0 | 155(57.6%) | 114 (42.4%) |
|  | High (≥126 mg/dl) | 121 | 31.0 | 94 (77.7%) | 27 (22.3%) |
| Total cholesterol | <200 mg/dl (Normal) | 276 | 70.8 | 169 (61.2%) | 107 (38.8%) |
|  | ≥200 mg/dl (High) | 114 | 29.2 | 80 (70.2%) | 34 (29.8%) |
| Triglyceride | Normal (<150 mg/dl) | 143 | 36.7 | 95 (66.4%) | 48 (33.6%) |
|  | High (≥150 mg/dl) | 247 | 63.3 | 154 (62.3%) | 93 (37.7%) |
| HDL-C | Normal (>60 mg/dl) | 203 | 52.1 | 98 (68.0%) | 32 (65.7%) |
|  | Low (≤60 mg/dl) | 187 | 47.9 | 111 (59.4%) | 46 (40.6%) |
| LDL-C | Normal (<130 mg/dl) | 325 | 83.3 | 196 (60.3%) | 129 (39.7%) |
|  | High (≥130 mg/dl) | 65 | 16.7 | 53 (81.5%) | (18.5%) |

### Prevalence of hypertension among diabetic patients

The overall prevalence of the prevalence of HTN among diabetic patients was 43.6% [CI: 39.0, 48.5].

### Factors independently associated with hypertension

The factors significantly associated with HTN on multivariable logistic regression analysis were increased age, family history of HTN, being single and being obese (Table 4).

**Table 4:**
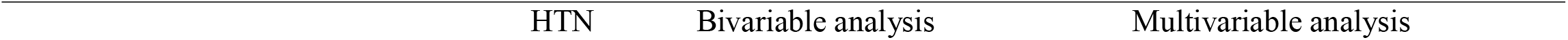
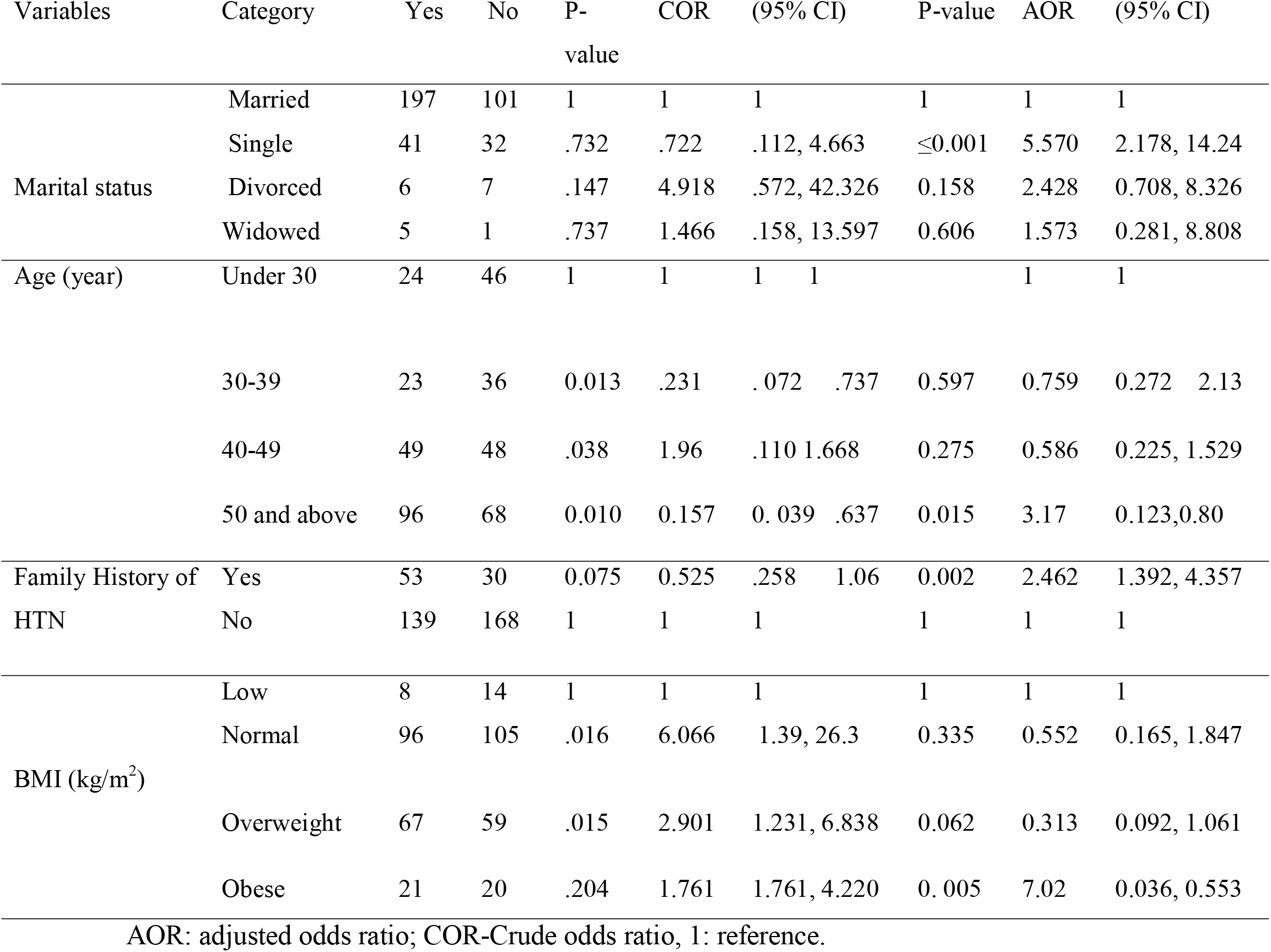
Factors associated with HTN among diabetic patients at public hospitals West Shewa, Ethiopia 2020

## Discussion

In this study, prevalence and determinants of HTN among DM patients were investigated in order to contribute to tackle the burden of the disease and its associated complications. The overall prevalence of HTN among DM patients on follow up at West Shoa Public hospitals was found to be 43.6% from the 390 participants [95%CI: 39.0, 48.5]. This result was in line with study done Pakistan 40.45% (23) and Taiwan 39% (24).

However, the finding of the current study is lower than a study conducted in Debre Tabor, Ethiopia (25) and Hosanna (55%), Ethiopia (26). Similarly, the prevalence of HTN in this study is higher than the findings reported in Southern Ethiopia at Sidama zone which reported 18.8%(27) and 25.6% in India (28). The possible reasons for difference with our finding might be due to differences in socio-demographic, study design, type of study population and sample size variation. The findings confirm the growing concern of HTN among DM patients as a public health problem and should be given attention.

Our study showed that patients with obesity have a higher risk of developing HTN than ones with normal BMI. This finding agreed with the findings of previous studies (29,30). The possible reason could be obese patients are at risk for developing arterial stiffness and endothelial dysfunction which increases blood pressure by rising renal tubular reabsorption, impairing pressure natriuretic, and causing volume expansion due to activation of the sympathetic nervous system and renin-angiotensin-aldosterone system (31).

In agreement with other studies, our study finding that revealed that age ≥ 50 years was independently associated with HTN among diabetic patients(32,33). The possible reason for this association might be due to with increasing age, there will be increasing stiffness of the aorta and aging is generally related with non-communicable diseases including HTN and decline in various physiological functions(34).

In agreement with one study, those participants who were single were 5.57 times more likely to develop HTN compared to married ones (35). But, other studies, have documented inconsistent results, hence further study is required to confirm this association (26,36). Possible justifications married individuals have better sleep, less stress, better moods and have a more healthy diet compared with never-married (37).

Finally, our study finding revealed that having family history of HTN was significantly associated with the occurrence of HTN among diabetes patients. The possible reason might be, even though the exact mechanism is not completely known, the probable explanation is that due to the fact that family members may share similar lifestyle and genetic factors. Besides, some studies have revealed that a positive family history of HTN is related with an initial rise in markers of inflammation in otherwise normal young normotensive individuals, likely conveying a predisposition to develop early atherothrombosis and rise risk of cardiovascular death(38,39).

## Conclusion

The study found majority of diabetes patients suffer from co-existing HTN. Furthermore the study pointed out that being obese, having family history of HTN, being single and older age was associated with HTN co-existing with DM. Thus, active search for early detection of HTN and related risk factors should be an important part of DM follow-up.

## Limitation of the study

First the cross-sectional design of this study was unable to identify a causal relationship. Second, since the study was hospital based, it may not be representative of the community and finally some questions were exposed for recall bias.

## Data Availability

The data sets used and/or analyzed during this study are available from the corresponding author upon reasonable request

## List of Abbreviations

AOR: Adjusted odds ratio
BMI: Body mass index
BSC: Bachelor of Science
CI: Confidence interval
COR: Crude odds ratio
DBP: Diastolic blood pressure
DM: Diabetes mellitus
ETB: Ethiopian birr
FBS: Fasting blood sugar
HTN: Hypertension
mmHG: millimeter of mercury
SBP: Systolic blood pressure
WHO: World health organization

## Declarations

### Ethical Approval

Ethical clearance was obtained from the academic research directorate of Ambo University, College of Health Science and Medicine, and the official letter of cooperation was written to the respective health facility heads and permission letters were obtained from the respective health facility heads. Then information was collected after written consent from each participant was obtained. Information was recorded anonymously and confidentiality and beneficence were assured throughout the study period.

## Acknowledgment

We would like to convey heartfelt gratitude for the study participants for their kind and unlimited cooperation, support and participation on the study. Last, but not least we want to acknowledge all persons who help us.

## Data Availability

The data of this study could be available on reasonable corresponding author request.

## Consent for publication

Not applicable

## Authors’ Contributions

DH conceived the idea, wrote the proposal, analyzed the data and drafted the paper, whereas DA participated in data collection, analyzed the data, and drafted the paper. Both authors read and approved the final manuscript data analysis and revised subsequent drafts of the paper.

